# A High-throughput Anti-SARS-CoV-2 IgG Testing Platform for COVID-19

**DOI:** 10.1101/2020.07.23.20160804

**Authors:** Jinwei Du, Eric Chu, Dayu Zhang, Chuanyi M. Lu, Aiguo Zhang, Michael Y. Sha

## Abstract

**Background:** Serology tests for detecting the antibodies to severe acute respiratory syndrome coronavirus 2 (SARS-CoV-2) can identify previous infection and help to confirm the presence of current infection.

**Objective:** The aim of this study was to evaluate the performances of a newly developed high throughput immunoassay for anti-SARS-CoV-2 IgG antibody detection.

**Results:** Clinical agreement studies were performed in 77 COVID-19 patient serum samples and 226 negative donor serum/plasma samples. Positive percent agreement (PPA) was 42.86% (95% CI: 9.90% to 81.59%), 55.56% (95% CI: 21.20% to 86.30%), and 96.72% (95% CI: 88.65% to 99.60%) for samples collected on 0-7 days, 8-14 days, and ≥15 days from symptom onset, respectively. Negative Percent Agreement (NPA) was 98.23% (95% CI: 95.53% to 99.52%). No cross-reactivity was observed to patient samples positive for IgG antibodies against the following pathogens: *HIV, HAV, HBV, RSV, CMV, EBV, Rubella, Influenza A*, and *Influenza B*. Hemoglobin (200 mg/dL), bilirubin (2 mg/dL) and EDTA (10 mM) showed no significant interfering effect on this assay.

**Conclusion:** An anti-SARS-CoV-2 IgG antibody assay with high sensitivity and specificity has been developed. With the high throughput, this assay will speed up the anti-SARS-CoV-2 IgG testing.

## 1. Introduction

A novel coronavirus, severe acute respiratory syndrome coronavirus 2 (SARS-CoV-2, previously provisionally named 2019 novel coronavirus or 2019-nCoV), has been identified as the source of a pneumonia disease (COVID-19) outbreak started in Wuhan China in late 2019 (1) and has caused a global pandemic. This virus is a single-stranded RNA virus with high sequence overlap to SARS-CoV. It contains nearly 29,900 nucleotides and has at least 14 open reading frames (ORFs): ORF1ab, spike (S), ORF3a, envelope (E), membrane (M), ORF8, and nucleocapsid (N) (2). A recent study demonstrated that both IgM and IgG antibodies were detectable 5 days after onset in all 39 patients with SARS-CoV-2 infection (3). The median day of seroconversion for both IgG and IgM was 13 days post symptom onset (4). Serological testing can help detect PCR-negative COVID-19 cases, especially for cases with high clinical suspicion but more than 7 days post symptom onset (5). Other clinical uses include epidemiologic survey of COVID-19 seroprevalence and identifying suitable subjects who are referred for donating convalescent plasma for potential therapeutic use. Serological testing may also help guide return-to-work decisions. Although many commercial testing products have been developed or are under development now, there are unmet needs for the sensitive and high throughput serological testing kits under the current global COVID-19 pandemic situation.

Herein, we reported the performance evaluation of the QuantiVirus™ anti-SARS-CoV-2 IgG test which is a two-step immunoassay using Luminex platform to detect anti-SARS-CoV-2 spike protein 1 (S1) receptor-binding domain (RBD) IgG antibody in human serum or plasma specimens. Validation of the test was conducted using COVID-19 negative and positive samples on both Luminex 200 and MAGPIX^®^ instruments. The test takes approximately 3 hours per run with a 96-well plate capable of testing 92 patient samples.

## 2. Methods

### 2.1 Instrumentation

According to the guidance issued by Centers from Disease Control (CDC) and the World Health Organization (WHO), all studies were conducted in a Biosafety Level 2 (BSL-2) cabinet when handling COVID-19 patient samples. The microplate shaker (PlexBio Co, Taiwan) was used for microplate shaking and incubation. Data acquisition was preformed on Luminex 200 and MAGPIX^®^ instruments (Luminex, Austin, TX).

### 2.2 Reagents and patient samples

The recombinant SARS-CoV-2 Spike protein 1 (RBD)-His containing 330-524 amino acids of Spike protein was produced from HEK293 suspension cells (ProMab Biotechnologies Inc, CA). SARS-CoV-2 Spike S1 Antibody (human chimeric, IgG isotype) was purchased from GenScript Biotech Corporation (Piscataway, NJ). Anti-SARS-CoV-2 Spike RBD monoclonal antibody (IgM isotype) was purchased from Creative Diagnostics (Shirley, NY). PE conjugated anti-human IgG Fc antibody was purchased from BioLegend (San Diego, CA). MagPlex Microsphere and xMAP^®^ Antibody Coupling (AbC) kit were purchased from Luminex (Austin, TX). Hemoglobin (human), bilirubin and EDTA were purchased from Sigma-Aldrich (St. Louis, MO).

Healthy donor EDTA K2 plasma samples were purchased from Golden West Biosolutions (Temecula, CA) in 2019 prior to the outbreak of COVID-19. COVID-19 negative EDTA K2 plasma samples were also obtained from University of Florida Department of Radiation Oncology in 2017. Healthy donor serum samples were purchased from Innovative Research, LLC (Plymouth, MN). COVID-19 patient serum samples were acquired from ProMedDx (Norton, MA) and University of California and VA Healthcare System.

Patient serum samples positive for IgG to *HBV/HCV/HIV/RSV* were purchased from Antibody Systems, Inc (Hurst, TX). Patient serum samples positive for IgG to *HAV/CMV/EBV/Rubella/Influenza B* were purchased from ProMedDx (Norton, MA). Patient serum samples positive for IgG to *Influenza A* were purchased from Dx Biosamples, LLC (San Diego, CA).

### 2.3 Assay procedure

Principle of the assay was shown in Figure 1. Recombinant spike protein 1 (S1) RBD was covalently coupled to the surface of MagPlex^®^ Microspheres (magnetic beads) via a carbodiimide linkage using xMAP^®^ Antibody Coupling (AbC) kit. S1 RBD protein coated magnetic beads and human specimens were mixed and incubated at room temperature for 1 hour. The IgG antibodies present in human specimens against S1 RBD protein (the antigen) will bind the coated magnetic beads. After washing, PE conjugated anti-human IgG antibody was added to the reaction mixture and incubated at room temperature for 0.5 hour. After washing, PE fluorescence of each well in a 96-well microplate was measured on Luminex 200 or MAGPIX^®^ instrument for Median Fluorescence Intensity (MFI). Interpretation of the testing results was performed by calculating the MFI ratio of each sample to the average MFI of two blank wells.

**Figure 1:**
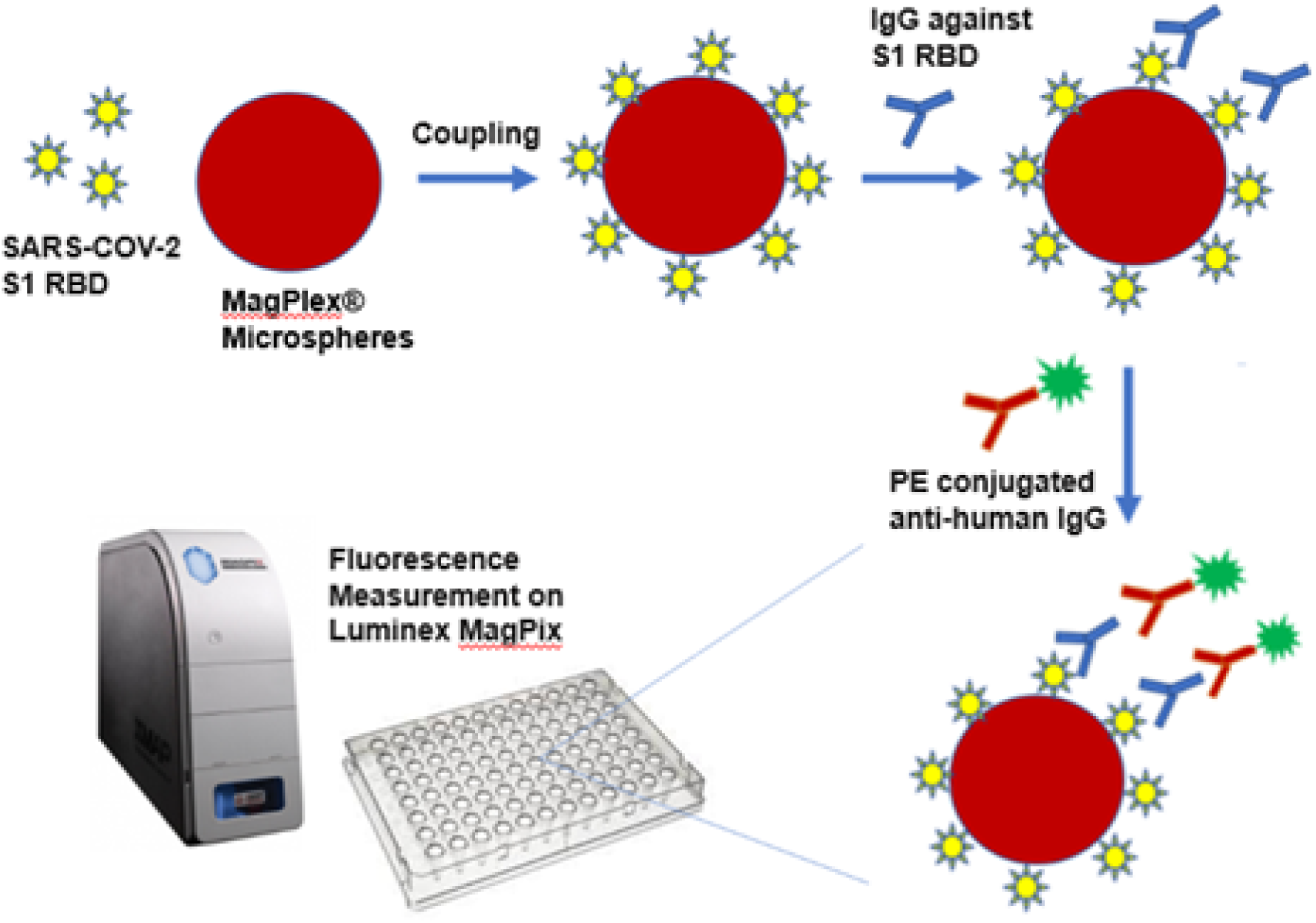
The high throughput Immunoassay for anti-SARS-CoV-2 IgG Detection.

### 2.4 Performance evaluation

To evaluate the clinical performance of the QuantiVirus™ Anti-SARS-CoV-2 IgG Test, 226 COVID-19 negative samples and 77 COVID-19 positive samples were tested and evaluated for NPA and PPA. Cross-reactivity was evaluated using serum or plasma samples which are positive for IgG antibodies to the following pathogens: *HIV, HAV, HBV, RSV, CMV EBV, Rubella, Influenza A*, and *Influenza B*. Class specificity was evaluated by spiking anti-SARS-CoV-2 RBD antibody IgG isotype and IgM isotype into a negative serum sample, respectively.

Within-run precision (repeatability) was evaluated by testing negative sample and positive sample in 21 or 24 replicates. Between-run precision was evaluated by testing negative sample and positive sample on five separate runs (3∼4 replicates per run).

For interference testing, hemoglobin (200 mg/dL), bilirubin (2 mg/dL) and EDTA (10mM) were spiked into four serum samples, respectively, and the MFI was compared with the control samples.

### 2.5 Statistical Analysis

For precision evaluations, coefficient of variation (CV) was calculated as the ratio of the standard deviation (SD) to the mean. For interference testing, the samples spiked with hemoglobin or EDTA were compared with the control samples by paired Student’s t-test with p≤0.05 defined as significantly different.

## 3. Results

### 3.1 Comparison between Luminex 200 and MAGPIX^®^

Unlike the Luminex 200, the MAGPIX system is not based on flow cytometry, but instead uses light-emitting diodes (LEDs) for excitation and a CCD camera for detection. Despite the difference in signal detection, the performance of the MAGPIX instrument has been shown to be comparable to the Luminex 200 instrument (6). To confirm this, we tested 5 COVID-19 negative samples and 4 COVID-19 positive samples with QuantiVirus™ Anti-SARS-CoV-2 IgG Test and performed data acquisition on both Luminex 200 and the MAGPIX^®^ instrument. As shown in Table 1, there was no significant difference in MFI values between Luminex 200 and MAGPIX (paired t-test, p=0.39) and the average concordance was 98% indicating high consistency between these two instruments.

**Table 1.**
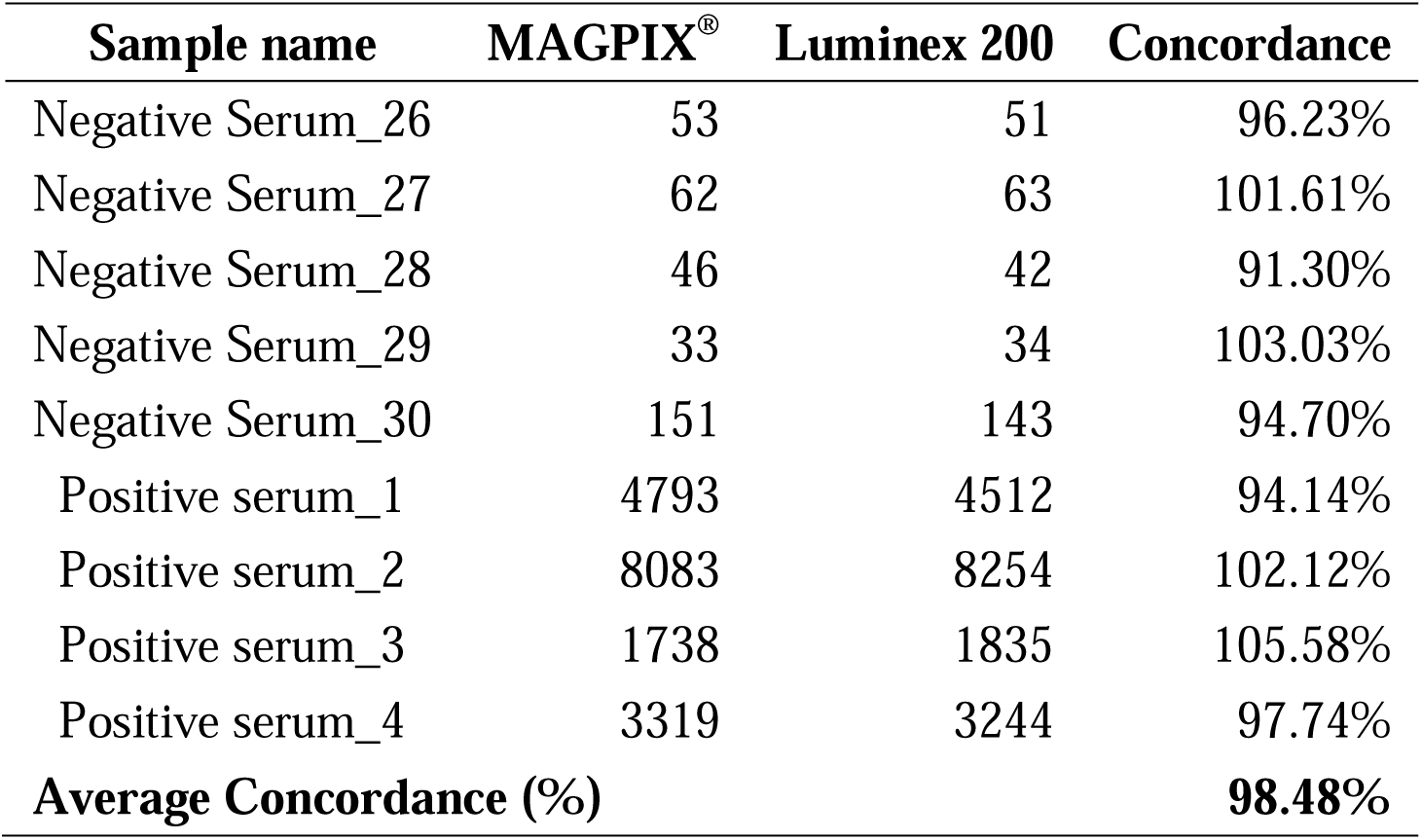
Fluorescence Signal (MFI) Comparison of Luminex 200 and MAGPIX.

### 3.2 Clinical Performance

Seventy-seven (77) serum samples collected at different times from individuals who tested positive with a RT-PCR method for SARS-CoV-2 infection were used in the evaluation of positive percent agreement (PPA). Two hundred and twenty-six (226) serum or EDTA plasma samples collected from healthy donors prior to the outbreak of COVID-19 were used in the evaluation of Negative Percent Agreement (NPA). As shown in Table 2, PPA was 42.86% (95% CI: 9.90% to 81.59%), 55.56% (95% CI: 21.20% to 86.30%), and 96.72% (88.65% to 99.60%) for samples collected on 0-7 days, 8-14 days, and ≥15 days from symptom onset, respectively, and NPA was 98.23% (95% CI: 95.53% to 99.52%).

**Table 2.** Positive Percent Agreement (PPA) and Negative Percent Agreement (NPA)

| Category | Days from Symptom Onset | Number of Samples | IgG Positive | IgG Negative | PPA and NPA (95% CI) |
| --- | --- | --- | --- | --- | --- |
| COVID-19 Positive | 0-7 days | 7 | 3 | 4 | PPA: 42.86% (9.90%~81.59%) |
|  | 8-14 days | 9 | 5 | 4 | PPA: 55.56% (21.20%~86.30%) |
|  | ≥15 days | 61 | 59 | 2 | PPA: 96.72% (88.65%~99.60%) |
| COVID-19 Negative | n/a | 226 | 4 | 222 | NPA: 98.23% (95.53%~99.52%) |

n/a: not applicable

Thirty (30) serum samples were also further evaluated by comparing to Abbott SARS-CoV-2 IgG antibody test which has been approved by FDA EUA. The results showed 100% concordance between the two assays (Supplementary Table 1).

In addition, 5 pairs of matched serum and EDTA plasma samples (i.e. collected from the same COVID-19 patients) were tested with QuantiVirus™ Anti-SARS-CoV-2 IgG Test and 100% concordance was observed. It indicates that serum and EDTA plasma is comparable for this test (Supplementary Table 2).

### 3.3 Cross-reactivity

Cross-reactivity of the QuantiVirus™ Anti-SARS-CoV-2 IgG Test was evaluated by using serum or plasma samples which are positive for IgG antibodies to the pathogens such as Influenza A or B (listed in Table 3). The result indicates that no cross-reactivity was found in any of the samples tested.

**Table 3.** Cross-reactivity Evaluation.

| **Category** | **Number of Samples Tested** | **Positive** | **Negative** |
| --- | --- | --- | --- |
| Human immunodeficiency virus (HIV) | 4 | 0 | 4 |
| Hepatitis A Virus (HAV) | 7 | 0 | 7 |
| Hepatitis B Virus (HBV) | 4 | 0 | 4 |
| Hepatitis C Virus (HCV) | 4 | 0 | 4 |
| Respiratory Syncytial Virus (RSV) | 5 | 0 | 5 |
| Influenza A | 5 | 0 | 5 |
| Influenza B | 13 | 0 | 13 |
| Cytomegalovirus (CMV) | 16 | 0 | 16 |
| Epstein-Barr Virus (EBV) | 13 | 0 | 13 |
| Rubella | 17 | 0 | 17 |

### 3.4 Interfering Substance

Hemoglobin (200 mg/dL) was spiked into four serum samples to test the potential interfering effect of high-level hemoglobin which might be present in hemolysis and other conditions. Bilirubin (2 mg/dL) was spiked into four serum samples to test the potential interfering effect of high-level bilirubin in the blood which might be caused by liver dysfunction such as hepatitis and cirrhosis. Lastly, EDTA (10mM) was spiked into four serum samples to test the potential interfering effect of EDTA which is the anticoagulant used in EDTA blood collection tubes.

As shown in Supplementary Table 3, the difference in the fluorescence signal (MFI) between the control samples and the samples spiked with hemoglobin, bilirubin or EDTA was all ≤ 11.5% which is acceptable to the test and no false negative or false positive results were observed. Therefore, hemoglobin, bilirubin and EDTA do not have significant interfering effect on QuantiVirus™ Anti-SARS-CoV-2 IgG Test at the tested concentrations.

### 3.5 Precision

Within-run precision (repeatability) was evaluated by testing negative sample and positive sample in 21 or 24 replicates. Between-run precision was evaluated by testing negative sample and positive sample on five separate runs (3∼4 replicates per run). As shown in Supplementary Table 4 and Table 5, the average CV% of within-run precision and between-run precision was 8.25% (from 4.71% to 11.74%) and 10.47% (from 5.0% to 13.4%), respectively.

**Table 4.** Class-specificity Test.

| **Sample** | **MFI-1** | **MFI-2** | **Average** |
| --- | --- | --- | --- |
| Negative serum sample | 19 | 23 | 21 |
| Negative serum sample with spiked human IgM isotype (5 µg/mL) | 18 | 18 | 18 |
| Negative serum sample with spiked human IgM isotype (0.5 µg/mL) | 17 | 18 | 17.5 |
| Negative serum sample with spiked human IgG isotype (2.5 µg/mL) | 824 | 840 | 832 |

### 3.6 Class Specificity

A study was performed to evaluate class specificity for QuantiVirus™ Anti-SARS-CoV-2 IgG Test. The human anti-SARS-CoV-2 Spike RBD antibody (IgG and IgM isotypes) was spiked into a negative serum sample respectively, and then tested by QuantiVirus™ Anti-SARS-CoV-2 IgG Test. A strong binding was observed for IgG isotype while no binding interaction was observed between the anti-human IgG antibody used in the QuantiVirusTM Anti-SARS-CoV-2 IgG Test and human IgM isotype, demonstrating class-specific reactivity only to human IgG isotype (Table 4).

### 3.7 Serum Heat-inactivation

Heat-inactivation is an effective means of destroying many types of virus and is used to protect the safety of laboratory workers exposed to blood and other body fluids while performing their jobs. Therefore, we tested if heat-inactivation will affect the results of QuantiVirus™ anti-SARS-CoV-2 IgG Test. As shown in Supplementary Table 6, heat-inactivation significantly increased the MFI for all the samples tested and one negative plasma sample became false positive, indicating that heat-inactivation is not suitable for the QuantiVirus™ anti-SARS-CoV-2 IgG Test.

## 4. Discussion

The RT-PCR tests designed to detect SARS-CoV-2 RNA have been the mainstay of testing for COVID-19 diagnosis and follow-up. However, serological testing should be helpful for detecting RT-PCR negative COVID-19 cases as well as asymptomatic infections (4). IgM can be an indicator of early stage infection, and IgG can be an indicator of current or prior infection. Besides epidemiological prevalence survey, IgG seropositivity can also be used to suggest the presence of post-infection immunity (7). Therefore, it is important to develop sensitive and specific serological testing for COVID-19.

Various methodologies for detecting anti-SARS-CoV-2 antibodies detection have been developed, including the traditional enzyme-linked immunosorbent assay (ELISA), immunochromatographic lateral flow assay, neutralization bioassay, and specific chemosensors (7). Development of high-throughput serology tests has also been a major focus of large diagnostics companies (8). Due to the specificity challenges associated with high false-positive rates, IgM may not play the primary role in COVID-19 antibody testing (9, 10). Therefore, we have developed the QuantiVirus™ anti-SARS-CoV-2 IgG test which can be run on Luminex platform for testing 92 samples per run within 3 hours. This high throughput assay has a sensitivity of 96.72% after 14 days from onset of symptoms and specificity of 98.23%. It can help clinical laboratories to further ramp up the COVID-19 diagnostics.

Recently, Cochrane Infectious Diseases Group assessed the diagnostic accuracy of anti-SARS-CoV-2 antibody tests using the Cochrane COVID-19 Study Register and the COVID-19 Living Evidence Database from the University of Bern (11). They found that the sensitivity of anti-SARS-CoV-2 IgG antibody test were highly variable with CLIA (94.6%), CGIA (87.3%) and ELISA (85.8%) all outperforming the lateral flow assay tests (76.0%). Panagiota et al. showed similar findings (12). Sensitivity also varies significantly with the time since symptom onset with average sensitivity 30.1% in the first week and 72.2% in the second week post-symptom onset (11).

FDA EUA approved Abbott SARS-CoV-2 IgG test on Abbott Architect platform reportedly has 100% sensitivity after 17 days from onset of symptoms and 100% specificity (13). The 100% concordance rate between the Abbott IgG test and the QuantiVirus™ anti-SARS-CoV-2 IgG test (Supplementary Table 1) indicates that the performance of our assay is comparable to that of the FDA EUA approved Abbott product.

Hettegger et al demonstrated that IgG profiles in plasma and saliva are highly similar for each individual and found the anti-HBV IgG antibody from saliva (14). We therefore tested saliva and serum from a COVID-19 patient. Although patient serum was detected for IgG, but no IgG signal from saliva (Supplementary Table 1, sample UCSF H). Since this was one patient sample testing, we cannot exclude the possibility of saliva as SARS-CoV-2 antibody detection. Further development effort is ongoing. Heat-inactivation is an effective means of destroying many types of virus to protect the safety of laboratory workers. It is also a standard procedure in diagnostic laboratories to conduct neutralization test for the purpose of inactivation of complement. Previous other virus studies suggested that serum heat inactivation and optimal dilution enhance WNV E-MIA sensitivity by eliminating the complement interference, thereby detecting low-titer anti-WNV antibodies during early and late phases of infection (15). Recently, two groups have reported that heat-inactivation of blood samples at 56 D for 30 minutes does not obviously affect the results of immunochromatography and chemiluminescent immunoassay for detection of SARS-COV-2 antibodies (16, 17). However, heat-inactivation cannot be used in fluorescence immunochromatography for SARS-CoV-2 antibody detection because negative samples became positive after heat-inactivation (16), which is consistent with our results that heat-inactivation significantly increased the MFI for all the samples tested and the negative plasma sample became false positive.

In conclusion, we have successfully developed a reliable high-throughput immunoassay for qualitative detection of anti-SARS-CoV-2 IgG antibody. The assay was validated with COVID19 positive samples as well as negative samples obtained from healthy donors on both Luminex 200 and MAGPIX^®^ instruments. We believe that this assay will help to determine the infection status of COVID19 and the true scope of the pandemic. It may be used to guide return-to-work decisions.

## Data Availability

all data from this manuscript are available for public

## Author contributions

J. Du, E. Chu and D. Zhang conducted the experiments. J. Du and M. Sha wrote the draft of the manuscript. M. Sha, CM. Lu and A. Zhang reviewed and edited the manuscript. All the authors read and approved the final manuscript.

## Declaration of competing interest

The authors declare that they have no conflicts of interest related the contents of this article.

## Notes

### Competing Interest Statement

The authors have declared no competing interest.

### Funding Statement

not application

### Author Declarations

health serum samples were purchased from Golden West Biosolutions (Temecula, CA) and Innovative Research, LLC (Plymouth, MN); COVID-19 patient serum samples were acquired from ProMedDx (Norton, MA)

## References

1. N. Zhu, D. Zhang, W. Wang, et al. A Novel Coronavirus from Patients with Pneumonia in China, 2019. N Engl J Med 382 (2020) 727–733.

2. M. Holshue, C. DeBolt, S. Lindquist, et al. First Case of 2019 Novel Coronavirus in the United States. N Engl J Med 382 (2020) 929–936.

3. M. Loeffelholz and Y. Tang. Laboratory diagnosis of emerging human coronavirus infections - the state of the art. Emerg Microbes Infect 9 (2020) 747–756.

4. Q. Long, B. Liu, H. Deng, et al. Antibody responses to SARS-CoV-2 in patients with COVID-19. Nat Med 26 (2020) 845–848

5. R. Patel, E. Babady, E. Theel, et al. Report from the American Society for Microbiology COVID-19 International Summit, 23 March 2020: Value of Diagnostic Testing for SARS-CoV-2/COVID-19. mBio 11 (2020) e00722–20

6. Comparison of Luminex® 200™ to MAGPIX® using the Poultry Serology Assay Technical Notes (011011-309-01). http://www.bioon.com/z/luminex/reference/poster-6.pdf

7. L. Carter, L. Garner, J. W Smoot, et al. Assay Techniques and Test Development for COVID-19 Diagnosis. ACS Cent Sci 6 (2020) 591–605

8. Serology testing for COVID-19. Johns Hopkins Center for Health Security. https://www.centerforhealthsecurity.org/resources/COVID-19/COVID-19-factsheets/200228-Serology-testing-COVID.pdf, accessed on 4/21/2020

9. M. Landry. Immunoglobulin M for Acute Infection: True or False? Clin Vaccine Immunol 23 (2016) 540–5

10. M. Bohn, G. Lippi, A. Molecular, et al. Molecular, serological, and biochemical diagnosis and monitoring of COVID-19: IFCC taskforce evaluation of the latest evidence. Clin Chem Lab Med 58 (2020) 1037–1052

11. J. Deeks, J. Dinnes, Y. Takwoingi, et al. Antibody tests for identification of current and past infection with SARS-CoV-2. Cochrane Database Syst Rev 6 (2020) CD013652

12. P. Kontou, G. Braliou, N. Dimou, et al. Antibody Tests in Detecting SARS-CoV-2 Infection: A Meta-Analysis. Diagnostics (Basel) 10(2020) 319

13. A. Bryan, G. Pepper, Mark. Wener, et al. Performance Characteristics of the Abbott Architect SARS-CoV-2 IgG Assay and Seroprevalence in Boise, Idaho. J Clin Microbiol (2020) 00941–20

14. P. Hettegger, J. Huber, K. Paßecker, et al. High similarity of IgG antibody profiles in blood and saliva opens opportunities for saliva-based serology. PLoS One 14 (2019) e0218456

15. M. Namekar, M. Kumar, M. O’Connell, et al. Effect of serum heat-inactivation and dilution on detection of anti-WNV antibodies in mice by West Nile virus E-protein microsphere immunoassay. PLoS One 7 (2012) e45851

16. X. Xue, C. Zhu, S. Huang, et al. Effect of heat inactivation of blood samples on the efficacy of three detection methods of SARS-CoV-2 antibodies. Nan Fang Yi Ke Da Xue Xue Bao 40 (2020) 316–320

17. X. Hu, R. Zhang, T. An, et al. Impact of heat-inactivation on the detection of SARS-CoV-2 IgM and IgG antibody by ELISA. Clin Chim Acta 509 (2020) 288–292

